# Investigating hsCRP as a clinical inflammation marker for human Bisphenol A food contamination offers protocol suggestions for conducting replicable, causal dietary intervention studies

**DOI:** 10.1101/2020.10.25.20212282

**Authors:** W. Lewis Perdue, Victor I. Reus, Rebecca L. Yeamans-Irwin

## Abstract

Dietary intervention studies thus far have failed to be replicable or causal. The results, therefore, have failed to provide clinicians and the general public with consistent and useful information on which to base reliable food-related health decisions. This is particularly relevant regarding plastic-derived chemicals (PDCs), such as Bisphenol A, now that the federal CLARITY-BPA program has failed to achieve scientific consensus. Investigators propose a novel human dietary protocol that is both replicable and causal, based upon BPA’s demonstrated inflammatory effects in humans. This first-of-a-kind dietary intervention study explores a potential causal relationship between human serum levels of BPA and High-Sensitivity C-Reactive Protein (hsCRP), a proven clinical indicator of inflammation. Investigators used the equivalent of a USDA-defined “typical diet” followed by a PDC-reduced diet to compare blood levels of hsCRP. This proof-of-concept investigation is the first to use an easily accessible, medically-accepted clinical laboratory test to directly measure human health effects of PDC reduction. Unexpected new complications discovered during the investigation indicate that these results may yet be inconclusive for direct causal relationship. However, the novel lessons and techniques developed as a result of those discoveries offer further specific and improved methods and best practices that can enable future dietary interventions to produce replicable, causal results.

## BACKGROUND

Exposure to environmental chemicals in the U.S. is widespread.^1^ As of June 2020, more than 86,000 chemicals were approved for use in the United States,^2^ and at least 4,000 of those are Plastic-Derived Chemicals (PDCs) present in food contact materials.^3,4,5^ PDCs such as Bisphenol A (BPA), phthalates, and other plastic derivations are present in approximately 97% of the U.S. population.^6,7^

Significantly, in a 10-year observational study of 3,883 adults in the United States, participants with higher urinary Bisphenol A levels were at a 49% greater risk for death from all causes.^8^

PDCs such as BPA are increasingly classified as endocrine disrupters, even in low-level concentrations.^7,9^

Human and animal studies have also identified PDCs compounds as contributors to cancer^10-21,^, cardiovascular disorders^12,22-29^, obesity^30-36^, type 2 diabetes^35,37-39^, metabolic syndrome^31-33,40,41^, neurological and behavioral disorders^42,43^ also including Alzheimer’s Disease^12,40,44-48^, as well as reproductive^13,49-56^, and developmental disorders^13,57-62^ and allergies^63-70^.

No scientific agreement exists about the health effects of most of these PDCs^71^. That includes BPA which, because of its ubiquity, has been singled out for special attention by the U.S. government as evidenced by the CLARITY-BPA^72^ program that was intended to resolve a contentious scientific divide on safety.

However, that divide still exists today as regulatory investigators have deemed current exposure levels as harmless^73-75^ while university-affiliated biomedical scientists continue to disagree^76^ and find BPA a public health threat.^77-79^

### Routes of Exposure

Exposure to BPA and other PDCs come from both dietary and non-dietary sources.^80^

BPA and other PDCs are found in household products such as detergents, cosmetics, lotions, and fragrances^81^, as well as in water bottles and baby bottles, thermal paper for printers, dental sealants, and medical devices including intravenous fluid and chemotherapy bags and tubing.^7,15,82-86^

Food and beverage packaging are substantial contributors to PDC physiological burdens.^7,87-91^ Consumers are exposed to many PDCs from leaching and migration of chemicals from plastics and other food contact materials.^7,88-96^

Exposure routes for food and beverages also include preservatives, flavorings, scents, texture enhancers, and coloring agents^5^, migration/leaching of chemicals from harvesting and processing^87^, as well as home food-handling which can accelerate migration through heating, microwaving, ultraviolet light exposure (including fluorescent lighting), and the contact of contaminated oils and alcohols with plastics.

Ultra-processed foods (UPFs) that have undergone substantial chemical or physical modification through manufacturing methods also have numerous avenues of PDC contamination due to residual contamination in additives, from additional contacts with plastic components, and from the addition of fats which facilitate the transfer of lipid-soluble polymer components into the UPFs.^98,99^

UPFs and PDCs are both associated with many non-communicable dietary-related diseases and syndromes including obesity,^100-107^ diabetes,^108-110^ cardiovascular disease,^111-113^ and cancer^10,18,114,115.^

### Could standard human clinical blood tests resolve the clarity issues with CLARITY?

As a result, the current CLARITY-BPA debate centers on experimental protocol flaws, contrasting interpretations of published science, confounding factors, sources of contamination and other fine points of scientific experimental design as well as practice and fundamental differences concerning the existence of non-monotonic behavior of substances present at very low levels.

Interpreting these factors by lay audiences can be all but impossible. Likewise, the current internecine arguments among scientists are unlikely to sway public, governmental or manufacturing opinion one way or another because:

1. There are no direct human health effects studies of BPA and other PDCs.
2. The CLARITY program has been criticized as irrelevant due to the failures of translating murine results to humans.^116^ Indeed, other published reports find that murine model results do not to translate accurately to humans as much as 92% of the time.^117^ Even pre-clinical research trials fail to be replicable^118^ from 51% to 89% of the time.^119-120^
3. Human dietary intervention studies, which might be able to shed light on the issue, have replication and scientific rigor flaws that prevent them from offering reliable clinical conclusions.^121,122^

### Enter the humble clinical blood test

Significantly, a source of reproducible, causally connected, and clinically valuable direct human data may lurk in standard laboratory blood profiles.

Because of the role of inflammation in many BPA-associated non-communicable diseases, High-Sensitivity C-Reactive Protein (hsCRP) may fill the need for clinical insights into numerous PDC-linked inflammation-linked conditions cardiovascular disease,^123-125,131,137^ Type 2 Diabetes,^126-127^ cancer,^126,129,137^Alzheimer’s Disease,^130, 132^ depression, and suicide,^133-136^ and auto-immune diseases^137^ including IBD,^138^ rheumatoid arthritis,^139^ and lupus.^140^

In addition, BPA has been found to activate the same NLRP3 inflammasome pathway activity^141^ implicated in “cytokine storms” which develop in research suspects along with bradykinin disorders^142^ as causes underpinning some of the most serious of COVID-19 cases.^143-145^

## OBJECTIVES

**Objective 1, validate marker:** Can hsCRP serve as a marker for BPA exposures?

**Objective 2, evaluate duration:** Is a short (e.g. six-day) trial long enough for an intensive BPA avoidance protocol to affect hsCRP outcome measurements?

**Objective 3, improve causality and replication:** Can replicability and causality of dietary intervention trials be increased by developing best practices and (for the first time) applying the discipline of standard laboratory practices to the sourcing, preparation, and serving of human food along with their complete data capture and reporting?

## METHODS

Given that the use of clinical blood tests as possible direct human health effects indicators of environmental chemical contamination is an unknown field, investigators felt that a small proof-of-concept trial (n-of-1) would be sufficient to test the validity of the concept and protocols before beginning a larger, more expensive study. N-of-1 investigations have found wide acceptance and success in biomedical science^146^ and are proposed as a main tactic in precision medicine^147^ and pharmaceutical research.

This n-of-1, six-day study (SSHS) was approved by the Committee on Human Research/IRB at the University of California San Francisco School of Medicine.^148^ The study consisted of two legs - a three-day “typical American diet” (Typical) with known sources of plastic contamination followed by a three-day intervention diet (Intervention) of foods with measurably reduced BPA contamination.

The study approved was based on a substantially revised protocol designed around a far more tightly controlled investigation. This investigation is intended to overcome inherent limitations on replicability in previously published studies that did not exercise investigator controls or data collection and other documentation involved with the sourcing of food, or its production and serving environment.

This investigation was conducted in a 400-square-foot professional kitchen supervised and directed by investigators in the same manner as a bench lab experiment. Central air with HEPA filters ventilated the kitchen which was also equipped with two standalone HEPA filter machines. A 1,200 cubic-feet/minute, four-centrifugal-fan exhaust hoods were used over an eight-burner natural gas stove. Two electric ovens were used for baking.

Plastic items including utensils, containers, and cookware with non-stick coatings were removed from the kitchen prior to trial onset. Food preparation took place on stainless steel countertops. Kitchen-ready scales were obtained and calibrated using brass gram weights.

Indoor kitchen air quality (via PM2.5 levels) were monitored via sensors from PurpleAir.Com and maintained at an Air Quality Index of 0. Test subject consumed meals in an adjoining space with identical environmental conditions including the absence of plastic.

### A. Menu Determination

The menu for each of the two diets (Typical and Intervention) was made as identical as feasible.

#### Typical

The “typical” American diet (i.e. PDC-contaminated diet) was drawn from the United States Department of Agriculture (USDA).^149^ Food for the typical diet was sourced from national brands available at a large national chain store (Safeway) to maximize availability.

#### Intervention

For the intervention diet, the menu from “typical American diet” was adapted to offer replicate food items with reduced or absent PDC contamination.

Intervention food was sourced in accordance with Appendix 2 of the revised protocol.^148^

The extensive requirements of the revised protocol included sourcing food close as possible to its actual production from a vendor capable of shipping nationally. Additionally, the producer:

- Either dry-farmed or used well water for irrigation,
- Did not irrigate with recycled wastewater or biosolids (sewage plant sludge)
- Adhered -- as a minimum -- to USDA organic standards.

Because milk is sourced locally or regionally, even by large chains, milk samples were analyzed via LC-MS/MS performed by Eurofins (https://www.eurofins.com/) to ensure the same or similar BPA quantification. Cheese products were selected from nationally available dairy brands whose milk scored below the limit of quantification (LOQ) in the LC-MS/MS tests.

Fish and seafood were excluded because the environmental pollution variability made it impossible for consistency in BPA exposure.

Carbon-filtered water was used for food preparation and to rinse before use of all dishes, utensils, pots, pans, and food-contact appliances. Multi-ingredient foods, such as a spaghetti and meatball frozen entrees, were “reverse engineered” from the dishes used in the “contamination” leg. Special attention was devoted to the health effects of micro- and nanoplastics, and to minimizing exposure to those particles of undeterminable composition.

### B. Food Preparation

After lengthy research, this study’s protocols were extensively revised^148^ to include the development of best practices that included the imposition of specific, rigorous scientific practices common to bench laboratory investigations. This included rigorous requirements for sourcing food and beverages, and the realization that a kitchen needed to be treated as a proper laboratory environment.

This required investigators to train and supervise all kitchen personnel to assure they were capable of precise measurements, and treating ingredients as reagents, recipes as procedures, appliances as equipment, and the paramount requirement to capture detailed and extensive data capture sufficient to allow replication of the trial under the exact same conditions.

In the kitchen, only glass, stainless steel, aluminum foil, and tight grain maple cutting boards were permitted for food preparation. 100% cotton dish towels were allowed; paper towels could only be used for cleanup. Vinyl gloves were used for all food preparation but contact with food products was designed to be minimal and incidental.

Intensive treatment of food in the revised protocol^148^ was designed to minimize BPA. For example, fresh organic vegetables were thoroughly rinsed or soaked in filtered water. Solid meat was first wiped with extra virgin olive oil obtained from a local mill which uses no plastic and packs the oil in glass. The meat was then scraped with a metal pastry divider and wiped dry again to remove lipid-solid PDCs such as BPA. Hamburger was prepared fresh using a grinder with all metal food contact surfaces.

No pre-prepared foods were allowed. Pasta and bread were baked using flour from a small mill that grinds wheat with no plastic contact obtained from small local organic, pesticide-free farms.

All measurements (including for liquids) were made by metric weight. All main ingredients were measured to the nearest gram, but small-weight items such as spices, herbs, other seasonings, and other items were measured to the nearest tenth of a gram.

Seasoning was with whole spices and fresh herbs with no PDC exposure grown for the purpose of this study by the investigators. When investigator-grown seasonings were not available, investigators specified organic whole spices (cinnamon sticks, whole nutmeg fruit, peppercorns) and hand ground them after wiping or rinsing with filtered water. Items with known or suspected plastic contamination (such as micro-plastic contamination of table salt^150^ were replaced with suitable reagents from Sigma-Aldrich.

### C. Blood Sampling of Study Participants

Two blood samples (one for hsCRP and one for BPA levels) were taken from each subject three times during the study: the first day to establish baseline; the last morning after the “typical” menu leg; and the last morning after the intervention leg.

All blood draws (hsCRP and BPA) were made at Sonoma Valley Hospital (SVH - an affiliate of UCSF).

Samples to be analyzed for hsCRP were drawn in light-green-topped tubes with standard concentrations of Li-heparin and centrifuged. Those were analyzed by the UCSF Parnassus campus medical laboratory.

The samples for BPA analysis were drawn by SVH using a vacutainer kit supplied by the lab of a UCSF researcher at the medical school’s Mt. Zion campus who had agreed to analyze the samples for BPA. The vacutainer kit was composed of polymers the researcher had found not to leach BPA or other PDC contaminants into the blood samples.

The samples were delivered as virgin whole blood to the UCSF researcher’s lab where they were centrifuged and frozen for LC-MS/MS analysis. Samples were hand-delivered within four hours of blood draws. To keep the high cost down, the samples were retained by the UCSF researcher’s lab and intended to be run as soon as a sufficient number of additional samples were in hand.

Originally, the hsCRP and BPA levels from the baseline, typical and intervention legs were to be compared to determine the degree of correlation. However, BPA sample analyses were not made available.

Accordingly, no hsCRP/BPA correlation was possible in this preliminary, proof-of-concept trial. However, correlations between hsCRP and the trial legs are presented as preliminary examination of the hypothesis.

## RESULTS

Levels of hsCRP decreased 21.4% from baseline to end of the Intervention leg: 1.1 mg/L from 1.4 mg/L. The results demonstrated a final percentage reduction in hsCRP parallel to (but smaller in magnitude) than that of a major NIH-funded dietary intervention by Hall, et al.^105^ published in 2019 approximately four months prior to our trial. (See Table 1)

**Table 1:**
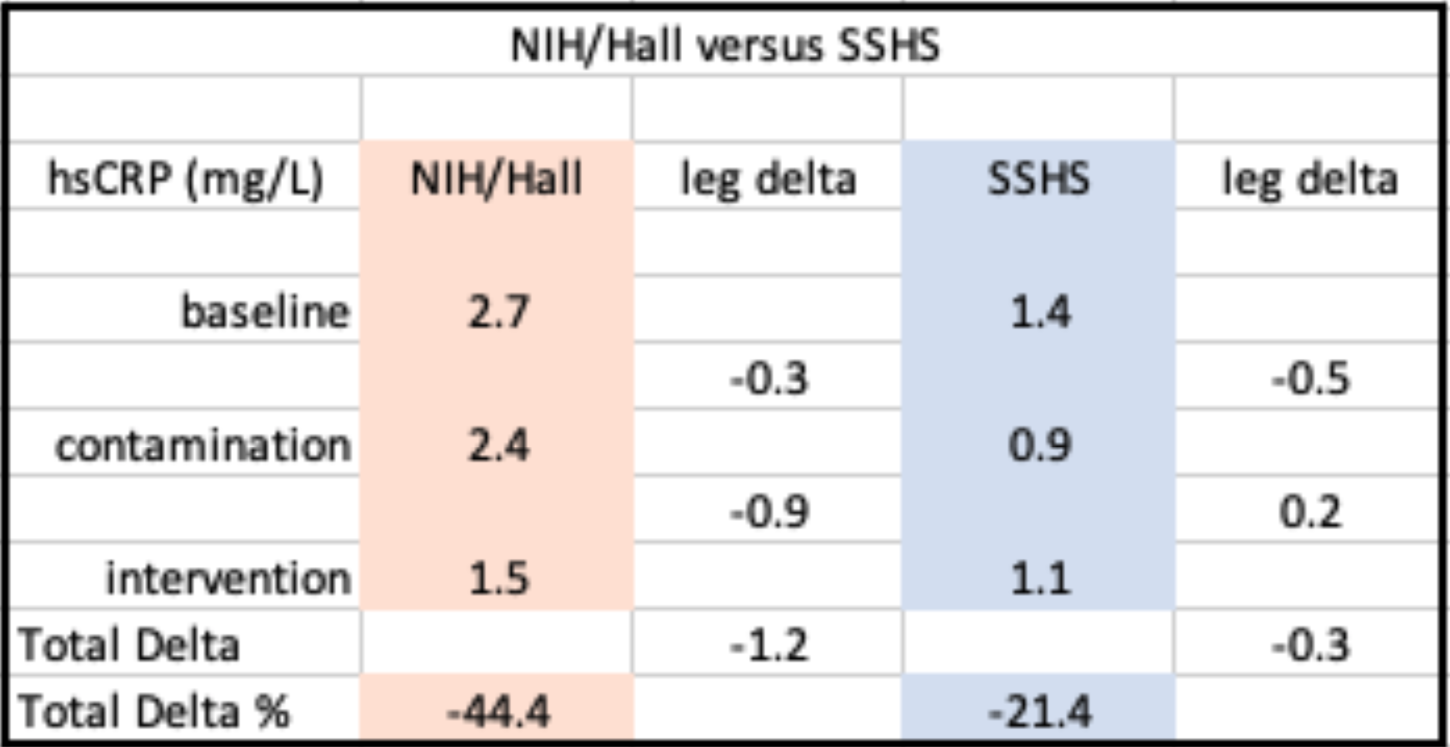
SSHS Study Results Versus Hall et al^105^.

## Discussion

This paper discusses a revised version^148^ of a study approved by UCSF-IRB on Nov. 15, 2014. Revisions were necessary because the original protocol was patterned after existing published dietary interventions,^73, 89^,^105, 150,151^ which lacked rigorous protocols capable of producing replicable results or credible causal relationships.^121,152^

Protocol revisions for this study were necessary because published dietary interventions involving BPA and other PDCs had relegated food sourcing and preparation to third parties – caterers or institutional kitchens – without investigator supervision, training in laboratory procedures, or proper data capture. A study without detailed data on food sourcing, preparation, procedures and serving environment cannot be replicated.

Most of this study’s protocol revisions involved developing best practices to bring human dietary interventions in line with the basic scientific rigor required of bench experiments. We believe those changes have made this investigation replicable.

However, replicable and causal considerations are not the same thing. A post-investigation review of Hall et al.,^105^ which was published four months before our trial, indicated that our investigation (as well as Hall et al.,) could not be causal because the “contamination legs” of both studies contained Ultra-Processed Foods which exert the same measurable biomarker influences as BPA. Complicating the situation further,there is the existence of BPA and phthalates in UPFs due to their more extensive plastic contact with processing and packaging.

For these reasons, neither study can conclude whether biomarker changes are causally due to BPA, UPFs, or both. This situation renders as irrelevant a further issue of whether nutritional differences in the two legs of the studies could influence the expression of relevant biomarkers.

Neither this study, nor Hall et al., succeeded in matching the nutritional values of both study legs for calories, total fat, saturated fat, trans fat, sugar, total carbohydrate, dietary fiber, or protein in order to account for possible health effects of those items. But even if investigators had been successful, these eight items are a small subset of the 26,000+ biochemicals found so far in foods,^153^ many of which are metabolically active and could influence health indicator biomarkers.

This knowledge dictates that establishing valid causal connections will require identical foods in both study legs, with food in the contamination leg dosed with an appropriate amount of the independent variable substance being evaluated for health effects.

The lower overall decrease in hsCRP in our study versus Hall et al., could be attributable to the shorter length of our trial. This may be due to the fact that BPA may have a longer half-life than previously thought.^154^ The lower decline in hsCRP levels in our study versus Hall et al. could have been caused by our shorter trial duration (six days rather than 28) which affects the release of BPA from adipose stores and/or its metabolism. Indeed, the minor increase of hsCRP in the second leg of the trial may indeed be due to the delayed release of BPA from the test subjects’ dietary changes.

An alternate explanation rests in the non-monotonic behavior of BPA, in which its effects become more powerful as the concentration falls into a specific range of influence.^155,156^

Non-monotonic behavior is counter-intuitive, but found in other compounds especially those that, like Bisphenol A, exhibit estrogenic effects active in the endocrine system. Breast-cancer treatment tamoxifen, for example, exhibits non-monotonic behavior and is most effective in tiny concentrations.^156^

A final difference between this trial and Hall et al., may be that our trial was interrupted by Northern California wildfires that dramatically increased PM2.5 particulate pollution, a notorious promoter of inflammation.^158^ This exposed the n-of-1 investigator/test to environmental PM2.5 pollution in order to deliver blood draw samples to two UCSF laboratories at the Parnassus and Mt. Zion campuses. Those exposures lasted for a minimum of 2.5 hours on three occasion. That level of exposure may account for the overall smaller decrease in hsCRP in our study versus Hall et al.

## Outcomes: Results of study objectives

### Objective 1, validate marker

#### Outcomes

The sourcing, preparation and serving protocols used in the intervention phase indicated that hsCRP levels were affected by the trial legs and may serve as a valid indicator. It is still unknown whether the lowered hsCRP level in our study can be a valid health indicator of human BPA contamination. not only because of confounding UPF factors, but also because the blood samples to be measured for BPA levels were reportedly misplaced by the LC/MS lab of the UCSF researcher who had agreed to process the samples.

However, this trial did reveal a pattern of hsCRP behavior consistent with Hall et al., which measured hsCRP as an indicator of inflammation in a trial of ultra-processed foods.

### Objective 2, evaluate duration

#### Outcomes

The study protocol’s projections of the metabolic pharmacokinetics of Bisphenol A and hsCRP point to the usefulness of the shortened trial length, although does not specifically confirm a test period as short as 6 days.

### Objective 3, improve causality and replication

This trial is replicable, but its results – like all other published human dietary interventions – does not illustrate acausal effect.

However, this study successfully developed best practices concerning the training and supervision of kitchen staff along with the sourcing, identification, preparation, and serving of foods, the use of scientific protocols, rigorous data capture, and the control of non-dietary exposures to PDCs that can lead to replicability and causality of subsequent dietary intervention trials involving Plastic-Derived Chemicals.

We did attempt to replicate the dietary portion of Hall et al,^105^ but our multiple requests to those investigators for useful information such as recipes, food sourcing never received a reply. The lack of that information along with that trial’s lack of use of unsupervised third-party food preparation make it incapable of replication. However, the use of a dormitory setting by Hall et al.^105^ is important in controlling the complicating conditions that could bias a dietary intervention study.

### Trials Replication and Causality

This proof-of-concept trial and associated research indicate that dietary intervention studies as a whole are inherently flawed and will not be replicable, causal, or lead to clinically relevant health recommendations or decisions unless they:

a. Apply basic scientific principles and record-keeping,
b. Conduct the study with human subjects,
c. Use exactly the same foods in both legs of the study,
d. Dose foods in the intervention stage using a single compound as an independent variable,
e. Eliminate non-food exposures and other confounding environmental and stress-related psychological confounders. This includes sequestering subjects in a disciplined but human-centered dormitory environment.

With proper attention to detail, these five requirements could possibly produce clinically useful methods for precision approaches to personalized dietary interventions.^147^

## Data Availability

Data, including supplemental materials, on request if not already posted and publicly available at https://stealthsyndromesstudy.com/

https://stealthsyndromesstudy.com/

## ACKNOWLEDGEMENTS

We would like to thank Dr. Alison Abritis Ph.D. for her helpful comments and invaluable discussions and suggestions.

This study was funded by the authors and the Center for Research on Environmental Chemicals in Humans, Sonoma, CA - https://crechcenter.org/

## SUPPLEMENTAL MATERIAL

Supplemental materials will be posted at https://stealthsyndromesstudy.com/ as they are formatted for upload.

